# Using Computer Vision and Artificial Intelligence to Track the Healing of Severe Burns

**DOI:** 10.1101/2022.12.15.22283475

**Authors:** Olivier Ethier, Hannah O. Chan, Mahla Abdolahnejad, Alexander Morzycki, Arsene Fansi Tchango, Rakesh Joshi, Joshua N. Wong, Collin Hong

## Abstract

Burn care management includes assessing the severity of burns accurately, especially distinguishing superficial partial thickness (SPT) burns from deep partial thickness (DPT) burns, in the context of providing definitive, downstream treatment. Moreover, the healing of the wound in the sub-acute care setting requires continuous tracking to avoid complications. Artificial intelligence (AI) and computer vision (CV) provide a unique opportunity to build low-cost and accessible tools to classify burn severity and track changes of wound parameters, both in the clinic by physicians and nurses, and asynchronously in the remote setting by the patient themselves. Wound assessments can be achieved by AI-CV using the principles of Image-Guided Therapy (IGT) using high-quality 2D colour images. Wound parameters can include wound 2D spatial dimension and the characterization of wound colour changes which demonstrates physiological changes such as presentation of eschar/necrotic tissue, pustulence, granulation tissue and scabbing. Here we present the development of AI-CV-based Skin Abnormality Tracking Algorithm (SATA) pipeline. Additionally we provide proof-of-concept results on a severe localized burn tracked for a 6-week period in clinic, and an additional 2-week period of home monitoring.

## 1 Introduction

According to a 2014 study, the total average healthcare cost per burn patient in high-income countries was $88,218 USD[9]. Inaccurate clinical assessments of the depth of the burn and non-standardized tracking of wound healing can add to the healthcare burden. Standardized characterization of burn depth and healing is paramount for burn case management by multidisciplinary healthcare teams. Current technologies like Laser Doppler Imaging(LDI) for ascertaining burn depths are in-accessible for most physicians and costly[6]. LDI uses laser light to scan the burn wound deep into the injured skin, including micro-circulation, thus providing information on the perfusion of the tissue as a surrogate for length of potential wound healing. In addition, there is no digital protocol that provides an easy means for quantitative, interval tracking of burn healing. The few related technologies that do exist[8], do not provide a mobile-friendly interface, that allows in situ care such as tracking a wound in clinic/community care, or for self-monitoring at home. Thus, the application of computer vision, AI and mobile device technology has garnered significant interest[16].

Current AI-enabled wound characterization methods can fit into two categories: standard computer vision and deep learning approaches. Standard computer vision methods sometimes require distinct hardware and usually need very constant conditions for accurate generalization[17][21][19]. Additionally, large, specific, and poorly accessible datasets are necessary for deep learning methods[25][7]. Among the standard computer vision approaches, segmentation of an object like a wound can be done by thresholding different combinations of pixel values extracted from the RGB and/or HSI colour representations[19]. Others use the mean shift algorithm[24] or histogram methods[11].

The most basic task of Convolutional Neural Networks (CNNs) is to classify images[12][23]. Deep neural networks like CNNs enable generalizations for unseen cases, making them more robust to heterogeneous backgrounds[13]. Using them to do segmentation is possible[15]. For instance, this has been achieved for foot ulcer images in the medical field[7][25]. Unfortunately, no well-curated datasets are available for burn wounds, which can be used for burn area segmentation.

As neural network models are also often seen as black boxes when it comes to decision-making, deconvolution[27] and guided backpropagation[22] are two methods which highlight the features of the input image that contributed most to the output results. However, these methods are not class discriminative. Gradient class activation maps (Grad-CAM), a generalization of CAM [28] is the state-of-the-art in terms of CNN decision-explanation[20]. Unfortunately, this feature saliency system returns very low-resolution attention maps that do not provide clinically actionable information. There is also the challenge of measuring the segmented area.

A simple medical protocol to measure wounds in real-world 2D values is to place an object of a known size next to the injury. Usually, a ruler or a grid is used [2][25]. Another method is to use a specific setup or hardware; For instance, Gaur et. al.[5] used a Time-Of-Flight camera for measuring and Filko et. al.[4] uses an RGB-D camera for full wound reconstruction.

Machine learning methods also exist for wound measurement. Carrion et al [1] used a deep learning framework to measure wounds, which required a well-defined frame around the wound as reference. Similarly, another study[26] used a support vector machine (SVM) for segmenting the wound and inferring its size, which used a dataset created by using a constant wound-to-camera distance.

When it comes to wound size and wound coloration during healing, studies show they are indeed correlated, but complementary [21]. There are multiple ways to process wound colour. For instance, some use raw RGB values to classify pixels in different tissue types: granulation or slough [19]. In contrast some studies introduce the HSI/HSV colour representation for analyzing wounds [10][18]. Apart from classifying into tissue types, a simpler approach is to report colour ratios instead. For example, Loizou et al. introduce a black-yellow-red and a red-yellow-black-white model, where each colour can be interpreted as a specific tissue type [14][3].

We introduce SATA (Skin Abnormality Tracking Algorithm), a suite of machine learning algorithms to characterize burns, which can also be applied to other skin wounds. This paper will discuss proof-of-concept results of how computer vision technology can be used to provide information regarding the severity of a wound, and the evolution of the wound, such as size and colour composition, even when the patient autonomously uploads images during their recovery phase from home. Through automation and standardization, the purpose of SATA is to be utilized as a decision aid by providing clinicians with objective data extracted from raw mobile camera images of wounds. This report also explores the feasibility of remote tracking of burns, where visualization data is provided to the data team by the patient who takes an active role in the at-home monitoring of the wound.

## 2 Methodology

### 2.1 Algorithms

SATA (Skin Abnormality Tracking Algorithm) comprises of three major components and functionalities. The components were coded in Python using python libraries, including OpenCV. The first component in SATA is wound segmentation, which first determines the location of the wound in an image. The second and third components are wound measurement and colour detection, which provides insights regarding the segmented wound. The segmentation mask and the raw image act as inputs to predict ratios of colours and wound area estimates, respectively (Fig.1). Since this method is intended as a tracking tool, we applied this pipeline to a time series of evolving wound images where each image is processed individually. The measured quantities are output as composite maps and stored in memory for time-series plotting.

**Figure 1:**
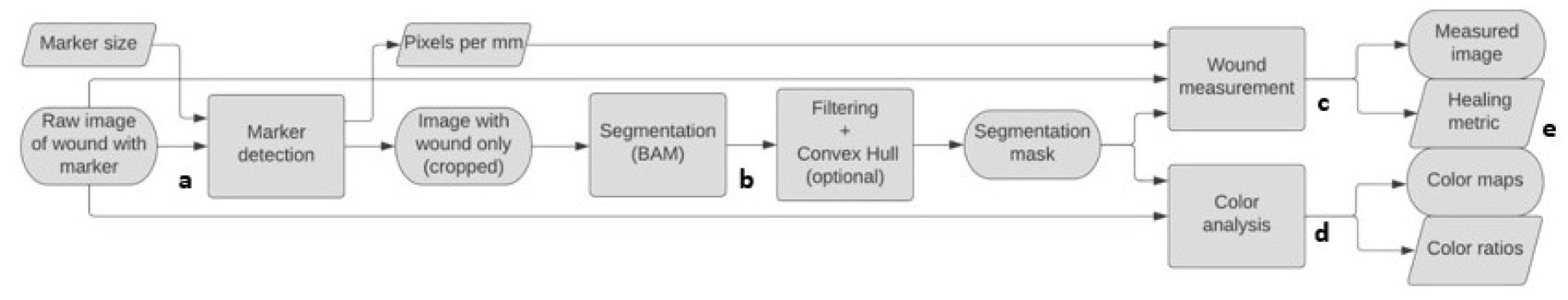
Skin Abnormality Tracking Algorithm (SATA) for Burn Injuries. The schematic depicts the SATA pipeline from an image input (a) to result outputs (e). Boxes illustrate scalar entities and processes, and ovals represent images. Once an image with a fiducial marker is uploaded, the system detects the fiducial marker for colour and size (a). This input image is processed to crop only the wound area and acts as input to the boundary attention mapper (BAM) component (b). The BAM output undergoes filtering, convex hull operations and masking, which act as inputs for (c) a wound measurement component, along with marker-based pixel/mm information, and (d) a colorimetric analysis component. The SATA pipeline culminates with outputs of a measured wound image, a healing metric, colour maps and a colorimetric analysis (e).

#### 2.1.1 Burn Segmentation: CNN

A CNN model based on a pretrained EfficientNetB7 [23] was used. The classifier head was altered to classify burn severity as superficial partial thickness (SPT) and deep partial thickness (DPT) in 2D images. Its structure was obtained after performing hyper parameter optimization, including adding a 4-layered perceptron, optimizing learning rates, regularization by optimizing drop-out variables, among other parameters. Increase in categorical validation accuracy and a decrease in validation loss were used to understand hyperparameter optimization in iterative rounds of training. The CNN model was trained on 1,285 2D colour images of burn patients, with an additional 273 images used as a validation set, from the Alberta Health System. The backgrounds in these images were removed by masking non-skin objects in the image field. Permission to use de-identified patient images was obtained under REBs from University of Alberta and the National Research Council, Canada.

#### 2.1.2 Burn Segmentation: BAM

Burn wound segmentation is conducted by the Boundary Attention Mapper (BAM) method, described elsewhere (Abdolahnejad et al, 2022 under revision). BAM builds on the principles of Grad-CAM to generate high-resolution saliency maps. Grad-CAM’s main incentive is to explain the predictions of a CNN classifier by highlighting the parts of the input images that contributed to the prediction. However, Grad-CAM outputs are typically coarser than the input images because they rely on feature maps late in the model’s architecture. BAM improves this resolution issue by finding feature maps early in the architecture as earlier feature maps have higher resolutions). The final BAM map is therefore based on selected early and late activation channel feature maps. We used the CNN described in the previous section to create the BAM maps.

The BAM output can have false-positive activated pixels surrounding the wound, especially with images taken at a later healing stage where a scar surrounds the wound. To make SATA more robust, we additionally process the BAM output by applying contour extraction and filtering them according to a custom criterion. This operation will be referred to as contour filtering as depicted in Figure 3. For each contour, the contour filtering algorithm iterates over threshold values and selects all contours with an area larger than it and fits a minimal-area rectangle over them. The criterion for selecting each set of contours (or each threshold) is the ratio of the selected contours’ total area over the box area. The set of contours maximizing this quantity is then selected as the wound contour(s). Generally, this results in selecting the single biggest contour in the list. However, if the segmentation function splits the region of interest in two or more neighbouring contours instead of one, the algorithm will select these contours altogether. Moreover, the BAM output may miss highlighting small parts of the region of interest around the edges. To cope with this, we apply a convex hull over the filtered contours. The resulting binary image will be referred to as the segmentation mask.

#### 2.1.3 Spatial Measurement of wound

As a reference for measurement, a round fiducial marker of a known diameter (here a coin with a diameter of 18.03 mm is used) was placed to the left of the wound. The fiducial marker in the images is identified by converting the image to grey scale, blurring it, extracting edges, and finding contours. The contours are then filtered to keep only the round ones that are bigger than a minimal size. The left most one is selected as the marker’s contour. However, slight angles between the skin surface and the camera’s view can make the marker appear tilted and thus more oval than round on an image. To address this issue, we fit a minimal-area rectangle over the oval fiducial marker contour and define a pixel-per-millimetre ratio by dividing its “largest diameter” (the longest axis of the oval) in pixels by the marker’s diameter in millimetres. Following this procedure guarantees that the pixel-per-millimetre rate is adequate in the axis of the longest diameter of the marker.

The image was first cropped and then fed to the segmentation function (to avoid the segmentation method from detecting the marker). Once a segmentation mask is computed, we use it with the original image to measure the wound area by squaring the pixels per millimetre value and multiplying it with the total amount of activated pixels in the segmentation mask.

Furthermore, we use the computed segmentation mask along with the original image to measure the wound area by squaring the pixel-per-millimetre ratio, denoted *ppm*, and multiplying it with the total amount of activated pixels in the segmentation mask. The wound area *A* can thus be calculated as *A* = *ppm*^2^ ∑ *M*_*i,j*_, where *M*_*i,j*_ is the pixel located at *i*^th^ row and *j*^th^ column of the segmentation mask and therefore is equal to 1 if that pixel is part of the wound and is equal to 0 otherwise. Thus, *i* ∈ [1, *H*] and *j* ∈ [1, *W* ] with *H* being the height and *W* the width of the image. In addition, a healing metric is directly derived from the measured wound area in the case where images of the same wound are available over time (i.e., time series of images). We define the biggest area registered in the series of wound images as the reference. All other images in the time series are attributed a remaining wound area percentage, from the ratio of its wound area over the reference.

#### 2.1.4 Wound colour analysis

To provide information about the wound composition, we use the Red-Yellow-Black-White model and provide information for these four colours for a clinical assessment. In this model, each colour of interest can be translated into a corresponding tissue type of clinical relevance. More precisely, red is associated with granulation and inflammation; yellow/white are associated with slough and scab; finally, black is associated with eschar and necrotic tissue.

The colour analysis is achieved by converting input images from the RGB colour representation to the HSL colour representation and applying thresholds for detecting the four colours of interest. The threshold values used in this study are shown in the Table 1. We compute colour ratios within the segmented region for each colour of interest and visualize the colour maps. Unlike the measurement process which provides an objective healing metric, the colour analysis provides qualitative information about the wound to act as a clinical decision aid in guiding the clinical assessment.

**Table 1:**
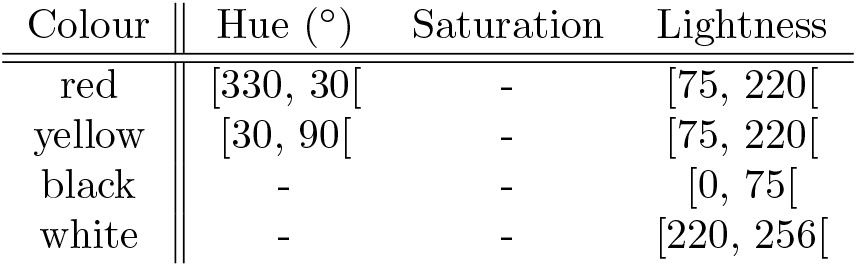
Threshold values in the HSL colour representation for detecting red, yellow, black and white. Hue is expressed in degrees ranging from 0 to 360. Saturation and lightness are expressed in pixel values ranging from 0 to 255 inclusively. For a pixel to be considered a given colour, its components must be in the intervals shown in this table. If no interval is shown for a certain component, then this component can have any value. For instance, a pixel will be considered white if its lightness is in the range [220, 256[, regardless of its hue and saturation values.

#### 2.1.5 Proof-of-Concept

As proof-of-concept, the healing of a localized burn on a patient in their 20s was tracked. The patient suffered a scald on the dorsum of the foot, resulting in a deep partial-thickness burn. Opting out of surgical intervention, the patient visited the clinic over a 6-week period for treatment with non-adhesive dressings and silver nitrate.

High-resolution images of the burn, with and without a fiducial marker (a Canadian dime coin was used as it is a common object), were captured with a smartphone camera (iPhone 8, iOS device) every 7-days. Images were taken under institutional or natural lighting and were used as algorithmic inputs. For continuity-of-care, the wound healing process was remotely tracked by asking the patient to send weekly images of their wound using their smartphone (Android device), both with and without any fiducial markers.

The patient consented to being remotely monitored for an additional 2 weeks, and electronically communicated images of the wound. The patient was instructed on how to capture these images through visual and written instructions. The images were to be captured in a format that would allow them to act as inputs to the first three algorithms, as well as capturing of stereoscopic 2D images, which will act as inputs for a disparity map-based algorithm being developed.

## 3 Results

### 3.1 Machine Learning components in SATA

A burn characterization pipeline (Figure 1) was created in Python using various OpenCV libraries for the wound measurement and colour analysis components. A CNN, based on the EfficientNetB7 architecture [23], was created that included an additional iteratively optimized MLP (Multi Layered Perceptron) layer (Figure 2 a). The model showed minimal over-training, although model training and training loss were not saturated (Figure 2 b and c). The CNN classifies 2D colour burn images as SPT versus DPT burns, with low false-positive and falsenegative rates (Figure 2 d). The f1-score of the model used for producing segmentation maps was 82% (Figure 2 e).

**Figure 2:**
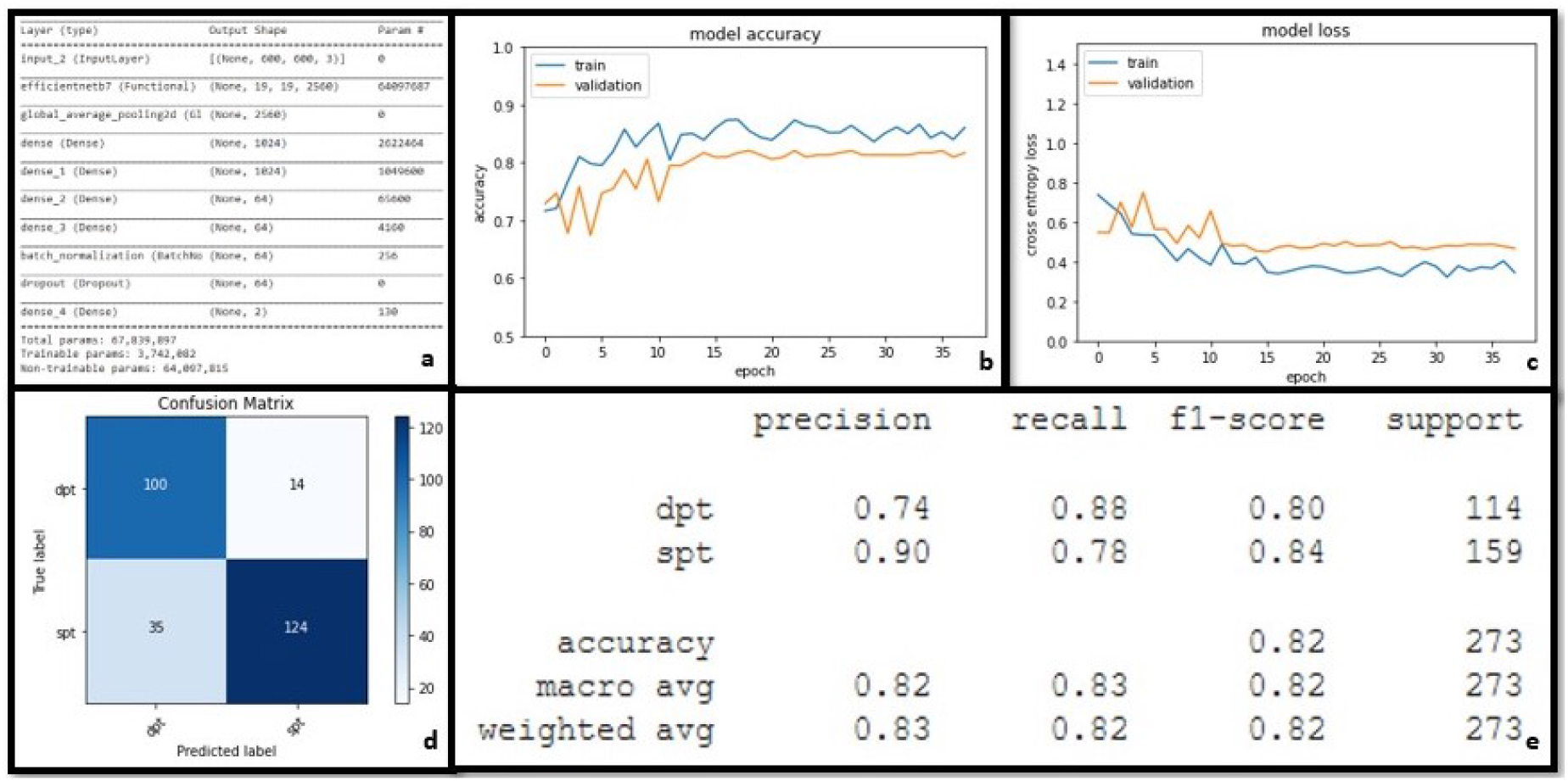
Neural Network metrics. The pre-trained EfficientNetB7 CNN (Convolutional Neural Network) architecture was further trained on images of two burn severity classes; SPT (superficial partial thickness) and DPT (deep partial thickness). A multilayered perceptron component composed of 4 dense layers, followed by a batch normalization layer and a dropout layer for regularization were added to the architecture (panel a). The plots of accuracy and loss over training epochs are displayed for the validation set and for the training set (panels b, c). A confusion matrix illustrates the error rates (including false positives and false negatives) from which specificity and sensitivities of the system can be extrapolated (panel d). Panel(e) reports the precision, recall, f-1 score, accuracy, and the number of validation images (support) associated with each class and on average (weighted averages and macro averages).

**Figure 3:**
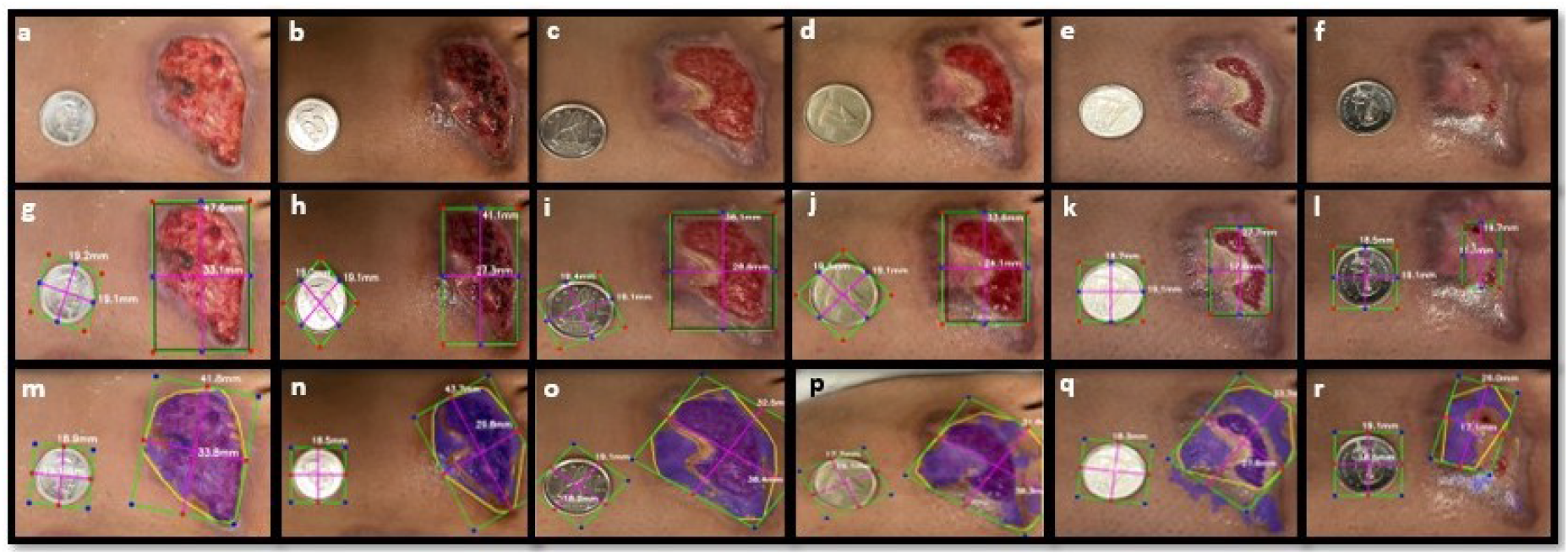
Burn wound 2-dimensional measurements. Images of a burn taken over a period of 35 days at 7-day intervals, using a mobile-device camera in clinic (panels a-f). Images are taken with a dime coin (18.03 mm diameter) as a fiducial marker for spatial references. The bounding boxes were digitally placed over the open wound on the images by clinicians as a ground truth, and computer vision is used to measure these bounding boxes (panels g-l). Contour masks were used to automatically mark the wound area (blue area), and the contour and convex hull application on the masks are outlined in yellow. For each mask contours the largest dimensions were measured (panels m-r).

This model’s select activation channel feature maps and Grad-CAM were used to create BAM maps to capture burn area boundaries. These BAM segmentation maps underwent contour-filtering (Figures 1 b, Algorithm 1) to fine-tune the BAM maps for automated wound measurement and wound colour analysis (Figures 1 c, d).

### 3.2 2D-wound measurement

A fiducial marker (a Canadian dime coin with a diameter of 18.03 mm) was used as a reference marker (Figures 1 a, 3 a-r) for 2-D wound measurement in millimetres (mm). Each image, taken 7 days apart, was taken with the fiducial marker placed near the injury site (Figures 3 a-f). These images either had a bounding box placed digitally over the wound area by a clinician, also referred to as manual-segmentation (Figures 3 g-l) or the BAM segmentation mask was used to automatically map the wound identified by the CNN (Figures 3 m-r).

Analysis of wound dimensions, either by manual-segmentation or BAM-generated masks, indicated a decrease of wound size over the 35-day period, both qualitatively and quantitatively (Figure 3). In the clinician segmented images, the open wound decreased from 47.6mm(height) x 33.1mm(width), with an error of ±1.17mm, to 19.7mm (height) x 11.1mm(width), with an error of ± 1.07mm (Figures 3 g-l). Similarly, for the BAM-based segmentation, the decrease in wound size was from 41.8mm x 33.8mm to 26.0mm x 17.1 mm (with a maximum error of ±1.17mm). When comparing the areas, the BAM-segmentation demonstrated a larger area for wound measurement which was clinically verified as areas of scabbing and hyperpigmentation around the wound.

Analyzing the two wound measurement methods quantitatively (Figure 4), we can see that the measurement done by the clinician shows a rapid decrease in size, especially in the initial stages (Figure 4 a), whereas the automated segmentation done by the BAM system, shows a more modest initial decrease in size, followed by a steeper decline (Figure 4 b). The two series of measurements (manual-segmentation and BAM-segmentation) are however highly correlated, with an average Pearson’s correlation coefficient of 0.87. This indicates that the BAM segmentation, with a higher sensitivity for colorations around the wound, provides an acceptable signal with respect to wound size, when compared to clinical assessment.

**Figure 4:**
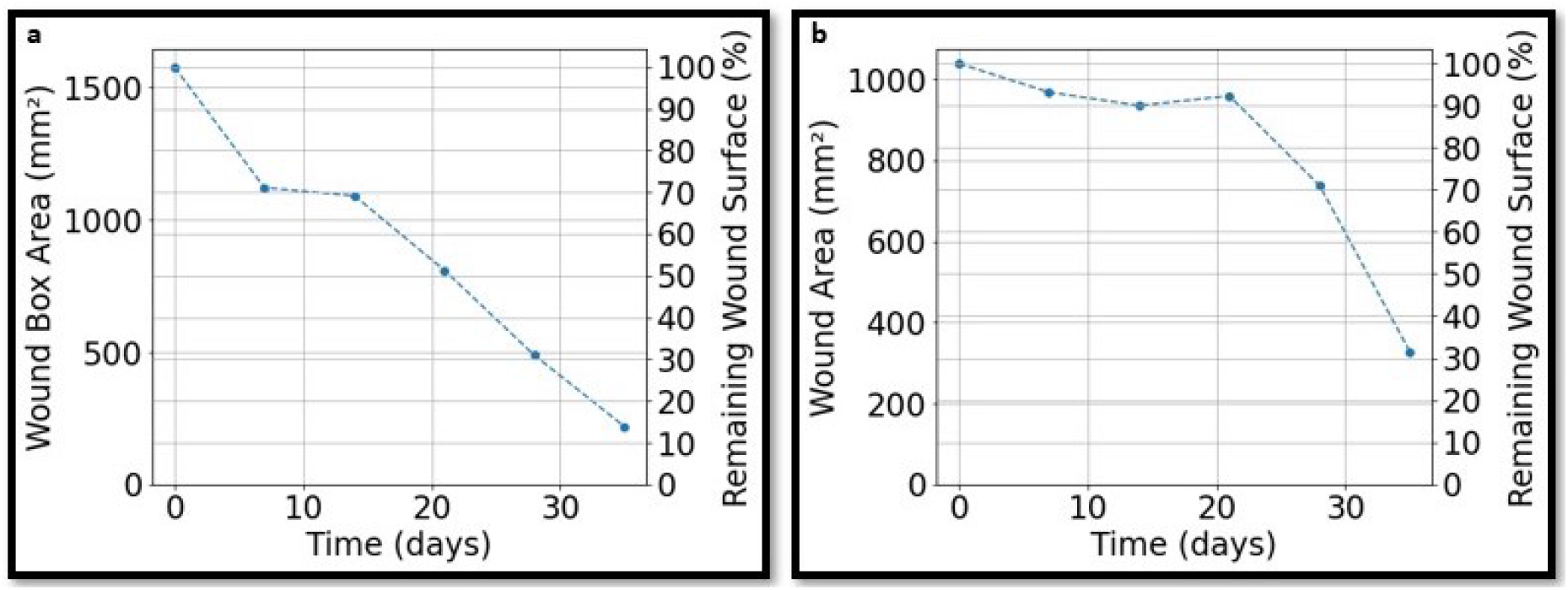
Burn wound measurement analysis. Quantitative analysis of the decrease of wound area, as mm^2^ and percentage of remaining wound area, using computer vision on clinical segmentation of open wound areas (a), and automated computer vision measurements using contour masks (b).

#### Algorithm 1

Skin Abnormality Contour Filtering. The steps and logic within the contour filtering algorithm to produce M¢ (a filtered binary mask) from M (a raw binary mask). The operators first identify and calculate the areas of all contours. The operations then iteratively select the contours based on certain area thresholds and use the selected contour for producing the mask.

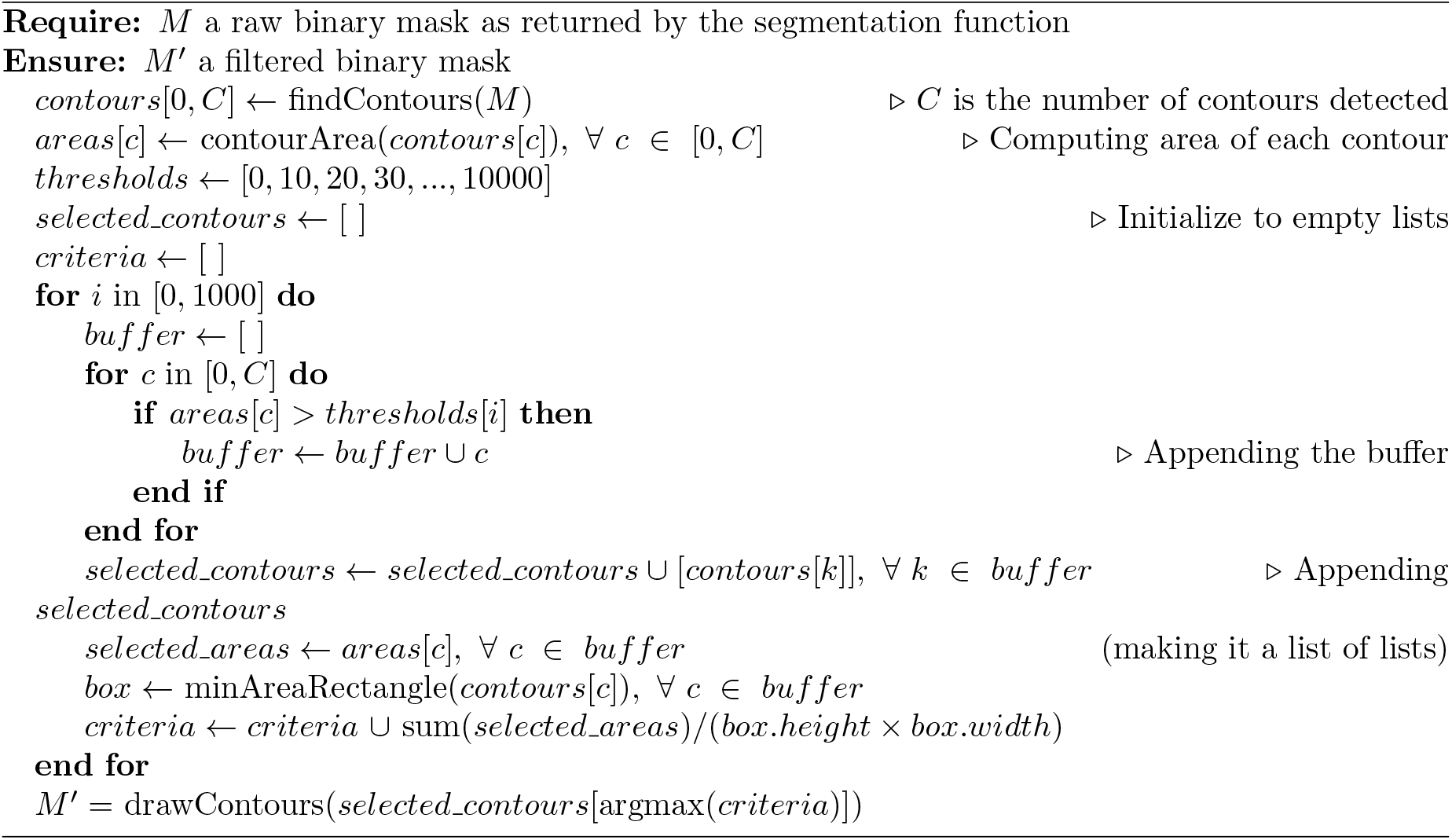

### 3.3 Tracking wound color

Qualitative colour maps of the wound over 35 days, and their quantitative analysis, for the burn injury are presented in Figures 5 and 6, respectively. The qualitative colour tracking results are illustrated in Figure 5 which depicts the tracked colours red (Figures 5 a-f), yellow (Figures 5 g-l), black (Figures 5 m-r), and white (Figures 5 s-x). We observe that red is the dominant colour within the wound, across the entire time series. Red can be interpreted as granulation tissue, especially since we observe cobblestone-like patterns in the burn injury (Figure 5). The ratio of detected red colour is greater than 90% of all colours tracked on average (Figure 6 a). This indicates that the healing tissue is the dominant tissue type according to the Red-Yellow-Black-White model. Interestingly, there is a peak in the black ratio at time step 2 (Day 14) (Figure 6 c) which would be indicative of eschar or necrosis. However, as the clinical assessment was normal healing progression, it was deep red/black blood being recognized by the algorithm. The algorithm also recognizes the paler left wound edge (scab) on Day 28 (Figures 5 k, 6 b). However, it is detected as yellow instead of white. Overall, the algorithmic assessment correlates highly with the clinical assessment and possesses the additional advantage of being more quantitative.

**Figure 5:**
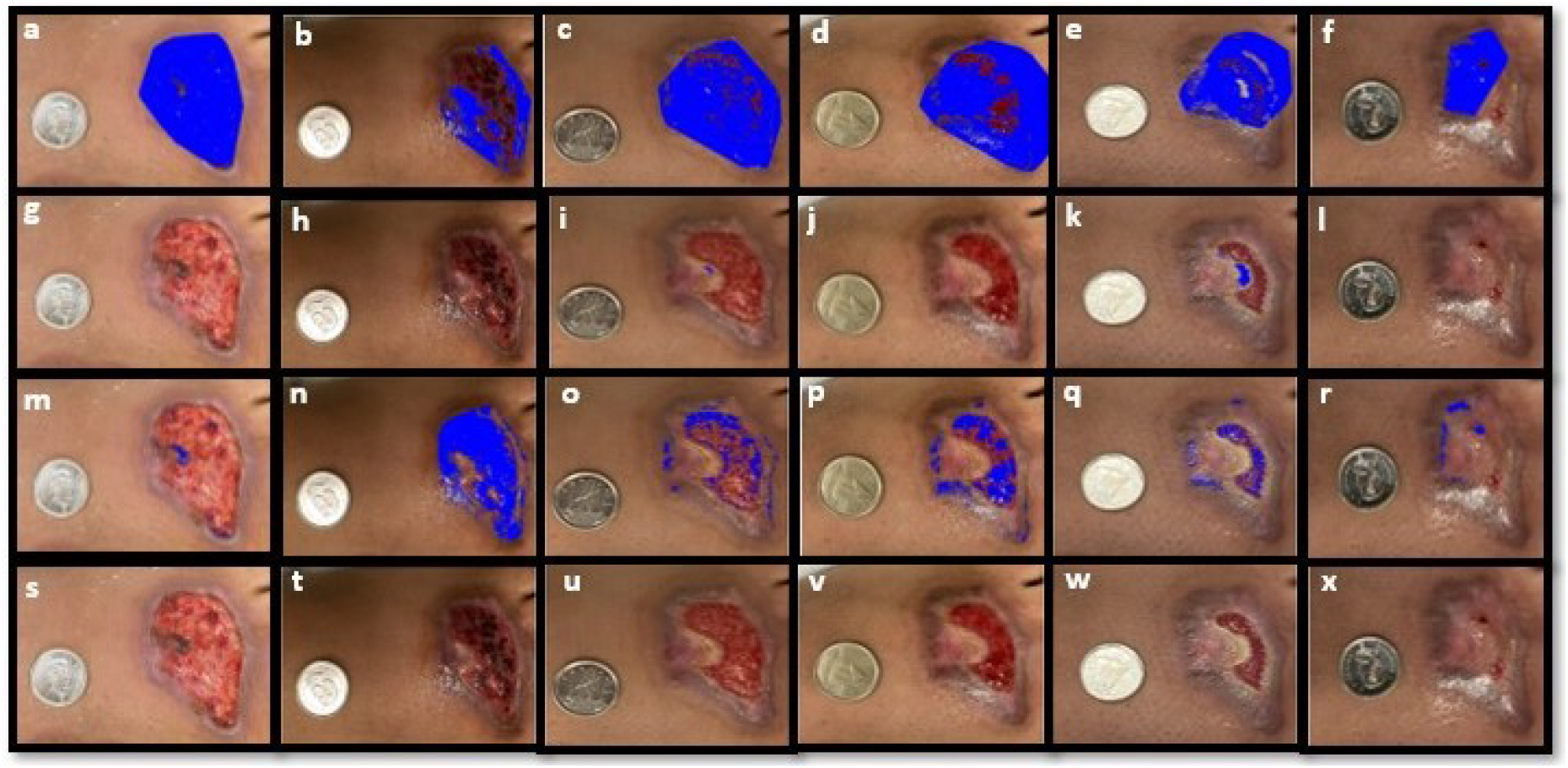
Burn wound colour tracking. Images of a burn taken over a period of 35 days at 7-day intervals, using a mobile-device camera in clinic, are tracked for measuring the red colour (i.e., granulation/inflammation) in the wound area, depicted by blue-pixeled area (panels a-f), for measuring the yellow colour (i.e., pustulence; panels g-l), and for measuring the black colour (i.e., eschar) (panels m-r). Lastly, epithelialization and scabbing are tracked via measuring the white pixels (panels s-x). All images were normalized for lighting and colour using the marker as a reference.

**Figure 6:**
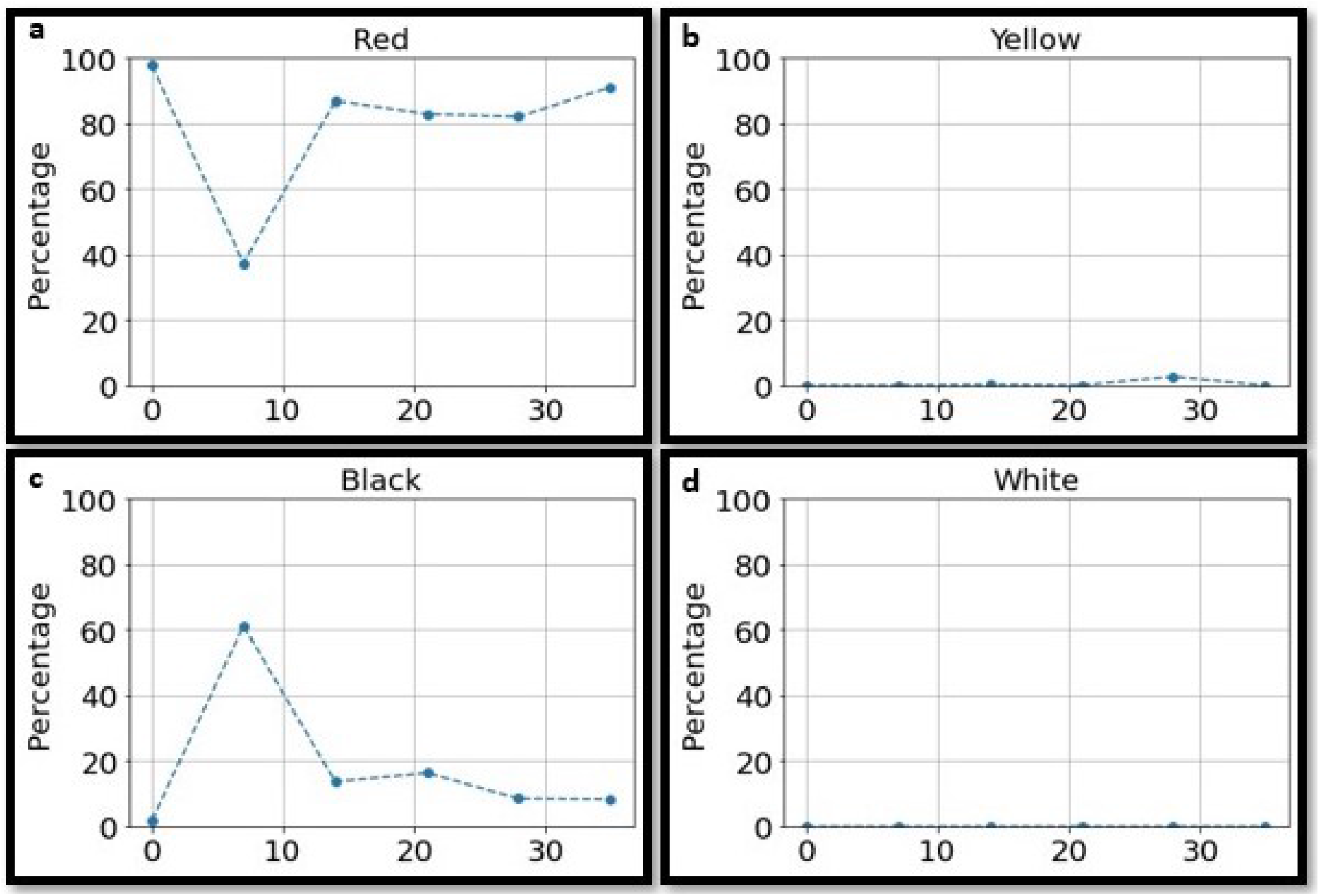
Burn wound colour analysis. Quantitative analysis of the changes in coloration of the burn wound (a-d), using computer-vision-aided colorimetric algorithm, over a 35-day period.

We continued with tracking of the burn patient’s healing process remotely (from home). The patient was instructed to take images of the wound on Days 42 and 49 (Figure 7) and electronically deposit them. Figures 87 a and b, illustrate the original images that were used to quantify the colors. Here we see that the red coloration is detected at a low level (20% range) and the white colour of the scabbing, is detected at 4 and 6% on Days 42 and 49, respectively (Figures 87 a, b, inset). We also see that the colour black is detected, due to the hyper-pigmentation around the healing wound, especially on Day 42 (Figure 7 a). We also see that manual segmentation and measurement of the burn injury (i.e., the pink healing tissue area; Figures 7 c, d) indicates a continued healing and an increase of epithelialization over the wound dermis.

**Figure 7:**
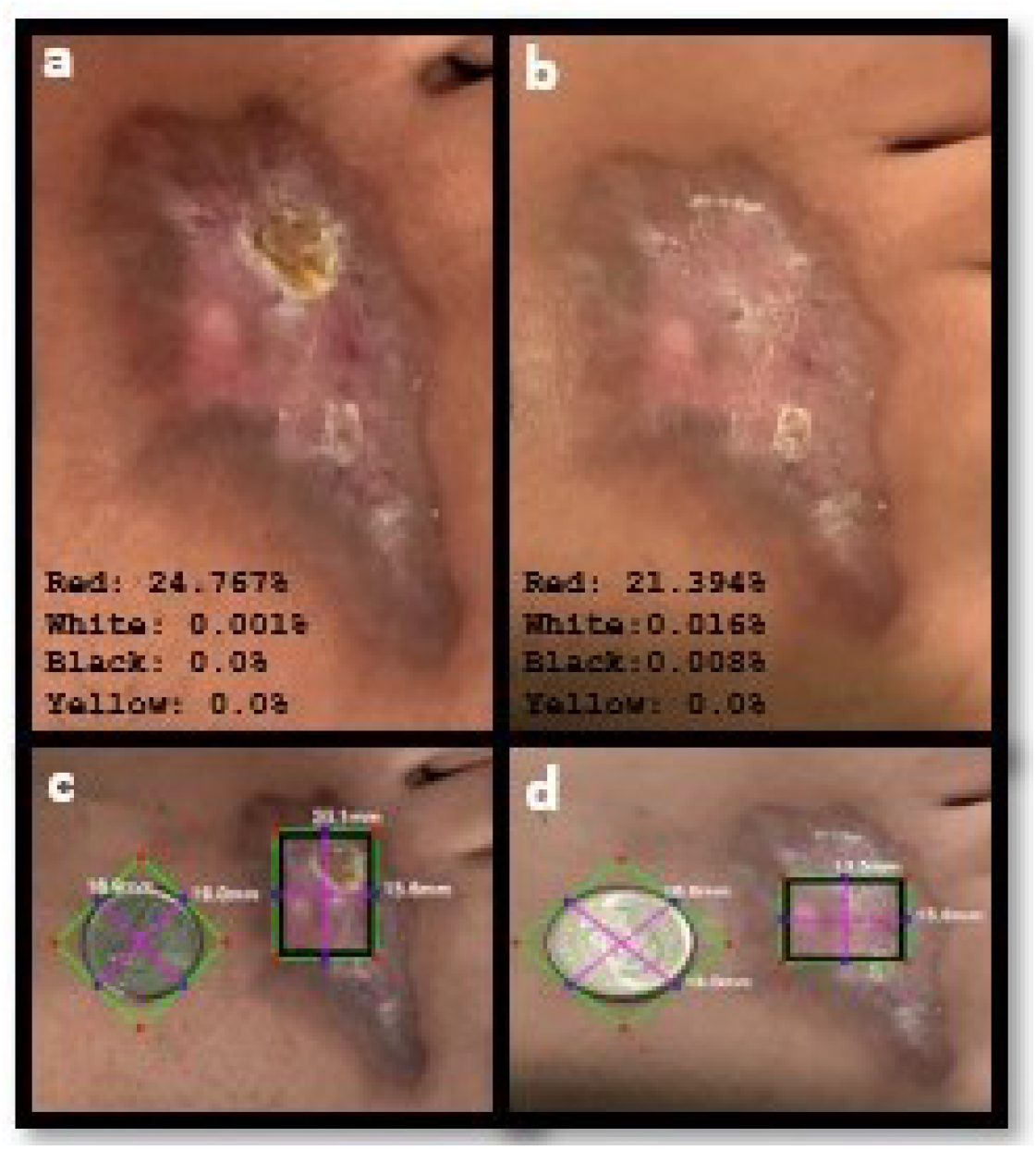
Remote wound tracking. The images taken by patient (at home) on days 42 (a) and 49 (b) of the burn wound were transmitted for SATA analysis. Inset values illustrate clinically relevant colour analysis. Panels (c) and (d) depict the clinician-segmented measurement of the wound, using a fiducial marker and computer vision.

## 4 Discussion

We present here the SATA (Skin Abnormality Tracking Algorithm) pipeline, a tracking tool for clinicians to follow the healing process of burned patients in clinic or remotely. This pipeline can be customized where any skin abnormality, from skin lesions to wounds, can be tracked in a standardized manner. The system is intended to work with minimal hardware requirements, namely a standard mobile-device camera and a round fiducial marker of a known diameter. We also demonstrate a novel method for wound segmentation termed the boundary attention mapper algorithm (BAM). BAM recycles deep convolutional neural network saliency maps, and improves the Grad-CAM mapping for finding contours.

Hence, the three main contributions of this study are:

1. Demonstrating the use of the Boundary Attention Mapper (BAM) as a wound segmentation tool based on deep learning. This is an improvements on the basic BAM which requires only a CNN classifier trained for the wound of interest and to be compatible with Grad-CAM.
2. Proposing a defined, but flexible pipeline for wound tracking using both wound area measurements and colour analysis with minimal hardware necessity.
3. Conducting a proof-of-concept for using SATA to provide standardized quantifications of the burn wound healing process in the clinic and remotely.

We demonstrate that a CNN can be reasonably accurate (over 80% in accuracy) when trained on a small dataset, for distinguishing a SPT burn from a DPT burn using colour skin burn images. This is a clinically relevant achievement as the delineation of these two severities of burn is critical for determining the next step in treatment, such as surgical intervention. The alternative is usually, an expensive and in-accessible assessment tool such as Laser Doppler Imaging to create a definitive assessment for clinical decision support.

The contour-attention map produced by BAM, which is fine-grained and dense, can be further processed using contour filtering and convex hull fitting. The system uses this segmentation map and the fiducial marker to provide information about both the area and the colour composition of the wound. We found that the automatically measured wound area has a Pearson’s correlation coefficient of 0.87 with the ground truth approach (clinical assessment) which consisted of manually putting a bounding box over the wound by an attending clinician. Also, the colour analysis results match our qualitative observations, but is more sensitive to slight changes in color. However, we see that further work is required for differentiating coagulated dark red blood from eschar/necrotic blackened tissue, which can be done by changing the range of values as reported in Table 1. This may also be true for differentiating the white-yellow spectrum but requires a larger dataset of wound injuries to test. We also found that the lighting of images plays a key role in accuracy of colour analysis of burn wounds, regardless of when they are taken under institutional lighting in a hospital setting or under natural light at home (data not shown).

This study provides proof-of-concept that algorithms if packaged in a mobile application with secure data sharing and connectivity to physicians, can bring the healing patient at home into the circle-of-care. Moreover, a burn classification system in a mobile app would also be invaluable to first responders, like EMS personnel responding to burn incidents, if provided with digitized Advanced Burn Life Support (ABLS) protocols. This aid will allow for more accurate initial assessments of a burn patient before reaching a care facility, by securely sending all patient data to downstream healthcare professionals. This includes burn specialists who can track the burn injury healing in the subacute care setting using the SATA pipeline if needed.

Future work includes testing the SATA pipeline on a larger set of burn time series images from a larger cohort, comparing results to current gold standards (such as LDI). SATA can also be tested on other skin abnormalities like ulcers and pathological wounds, experimenting with other segmentation approaches, and investigations of pixel clustering or classification from colour information. We hope this work will accelerate the development of pocket- and tele-medicine in burn care management.

## Data Availability

All data produced in the present work are contained in the manuscript

